# Evaluating the causal relationships between urate, blood pressure, and kidney function in the general population: a two-sample Mendelian Randomization study

**DOI:** 10.1101/2024.04.25.24306305

**Authors:** Haotian Tang, Venexia M Walker, Tom R Gaunt

## Abstract

**Background:** Associations between blood urate levels, blood pressure (BP), and kidney function have previously been reported in observational studies. However, causal inference between these three traits is challenging due to potentially bidirectional relationships. Method: We applied bidirectional univariable Mendelian randomization (UVMR) to assess the causal relationships between urate levels, BP, and kidney function, proxied by estimated glomerular filtration rate (eGFR), using genetic associations from UK Biobank and CKDGen. We performed multivariable MR (MVMR) to assess the independent effects of urate and BP on eGFR. Effect estimates are presented as standard deviation (SD) change in outcome (95% confidence interval) per SD increase in exposure.

**Results:** The UVMR analysis showed a bidirectional causal effect between urate and eGFR [urate on log(eGFR): beta=-0.10 (−0.22 to 0.02); log(eGFR) on urate: beta=−0.11 (−0.17 to −0.04)]. We also found bidirectional causal effects between urate and SBP [urate on SBP: beta=0.08 (0.04 to 0.11); SBP on urate: beta=0.13 (0.08 to 0.18)] and urate and DBP [urate on DBP: beta=0.09 (0.05 to 0.14); DBP on urate: beta=0.13 (0.08 to 0.18)]. However, there was weak evidence of a causal effect between BP and eGFR. MVMR results suggested the causal effect of urate on eGFR was independent of BP.

**Conclusion:** Our results provide evidence for bidirectional causal effects between urate and both eGFR and BP, suggesting urate control as a potential intervention to reduce BP and decline in kidney function in the general population, but little evidence of a causal relationship between BP and eGFR.

## Introduction

Hyperuricemia, defined as an elevated urate level in the serum, is a common disorder affecting about 20.1% of people in the United States [1]. Hyperuricemia is associated with several different disorders, including gout, hypertension, and chronic kidney disease (CKD) [2]. CKD is an irreversible and progressive disease, which can lead to end-stage renal disease requiring expensive treatments, such as dialysis or kidney transplantation. Elevated urate levels have been associated with a substantial positive risk of CKD in numerous epidemiological studies, and are a potential risk factor for the development and progression of renal disease in the general population [3–6]. However, urate, as the byproduct of purine metabolism, is primarily excreted via the kidneys. The natural relationship between urate and kidney function makes it difficult to identify whether the association between urate and CKD is causal, and if so, in which direction.

It has also been reported that higher urate levels are associated with an elevated risk of hypertension [7–9]. Primary hypertension patients commonly have hyperuricemia, which is more prevalent in patients with accelerated hypertension [10]. Hypertension and CKD are interlinked and represent huge global public health burdens, affecting around 31% [11] and 10% [12] of adults respectively. Renal function deteriorates with sustained hypertension, and blood pressure (BP) regulation deteriorates with progressive renal function loss [13]. Furthermore, the mouse uricase-knockout model has indicated higher urate levels affect the progression of hypertension and reduce kidney function [14]. Understanding the causal relationships between serum urate, BP, and kidney function might help reveal the underlying pathophysiological mechanisms and provide new evidence for clinical and lifestyle intervention.

Mendelian randomization (MR) can address some of the limitations of observational research including confounding and reserve causation, by using genetic variants as instrumental variables for an exposure to estimate causal effects on an outcome. MR relies upon the principle that alleles randomly segregate from parents to offspring, according to Mendel’s Laws of Inheritance. Consequently, it is unlikely that offspring genotypes will be linked to population confounders, such as behavioral and environmental factors. Furthermore, issues with reverse causation are avoided because germline genetic variants are established at conception and temporally precede the risk factors being researched [15, 16].

This study aimed to assess the causal relationship between urate, BP, and kidney function [proxied by creatinine-based estimated glomerular filtration rate (eGFR)] in the general population by conducting bidirectional univariable MR (UVMR) and multivariable MR (MVMR) analyses. In addition, we examined the effect of urate on hypertension at different points during the life course by estimating the effects of urate on early- and late-onset hypertension.

## Methods and materials

### Data source

We conducted genome-wide association studies (GWAS) in UK Biobank (UKB) (UKB project 15825), to obtain summary statistics for urate, systolic BP (SBP), diastolic BP (DBP), early-onset hypertension, late-onset hypertension, and overall hypertension [17]. The details of how we conducted GWAS are in Supplementary Materials. We also obtained summary statistics of both urate [18] and eGFR [19, 20], from the GWAS of European-ancestry participants carried out by the Chronic Kidney Disease Genetics Consortium (the CKDGen Consortium). We differentiate between the urate datasets from CKDGen and UKB GWAS by labeling them as “Urate (CKD-Gen)” [18] and “Urate (UKB)”, and distinguish between the two eGFR datasets from CKDGen as “eGFR (CKDGen2016)” [20] and “eGFR (CKDGen2019)” [19].

### Instrument Selection

We selected instruments robustly associated with each phenotype at genome-wide significance (p < 5e-8). Next, we used the ieugwasr R package to perform linkage disequilibrium clumping with a window of 10,000kb and a maximum *r*^2^ threshold of 0.001 to select independent variants [21]. We calculated F-statistics to evaluate the strength of the genetic instruments for each UVMR analysis [22, 23], using the TwoSampleMR R package [24]. F-statistics are typically interpreted using an arbitrary threshold of 10 as an indicator for a strong instrument.

### Main analyses

#### UVMR

To estimate the causal effects between urate, BP, and eGFR, we systematically conducted pairwise bidirectional two-sample UVMR analyses using the IVW method. We also examined the causal effects of urate on early-onset, late-onset, and overall hypertension using UVMR. Both urate and eGFR GWAS summary statistics were downloaded from the CKDGen consortium (http://ckdgen.imbi.uni-freiburg.de/) [19]. GWAS summary statistics for SBP and DBP were obtained by conducting novel GWAS in UKB [17]. All GWAS details and UVMR can be found in Table 2 and Table 1. All analyses are presented on the SD scale (see Supplementary Materials).

**Table 1:**
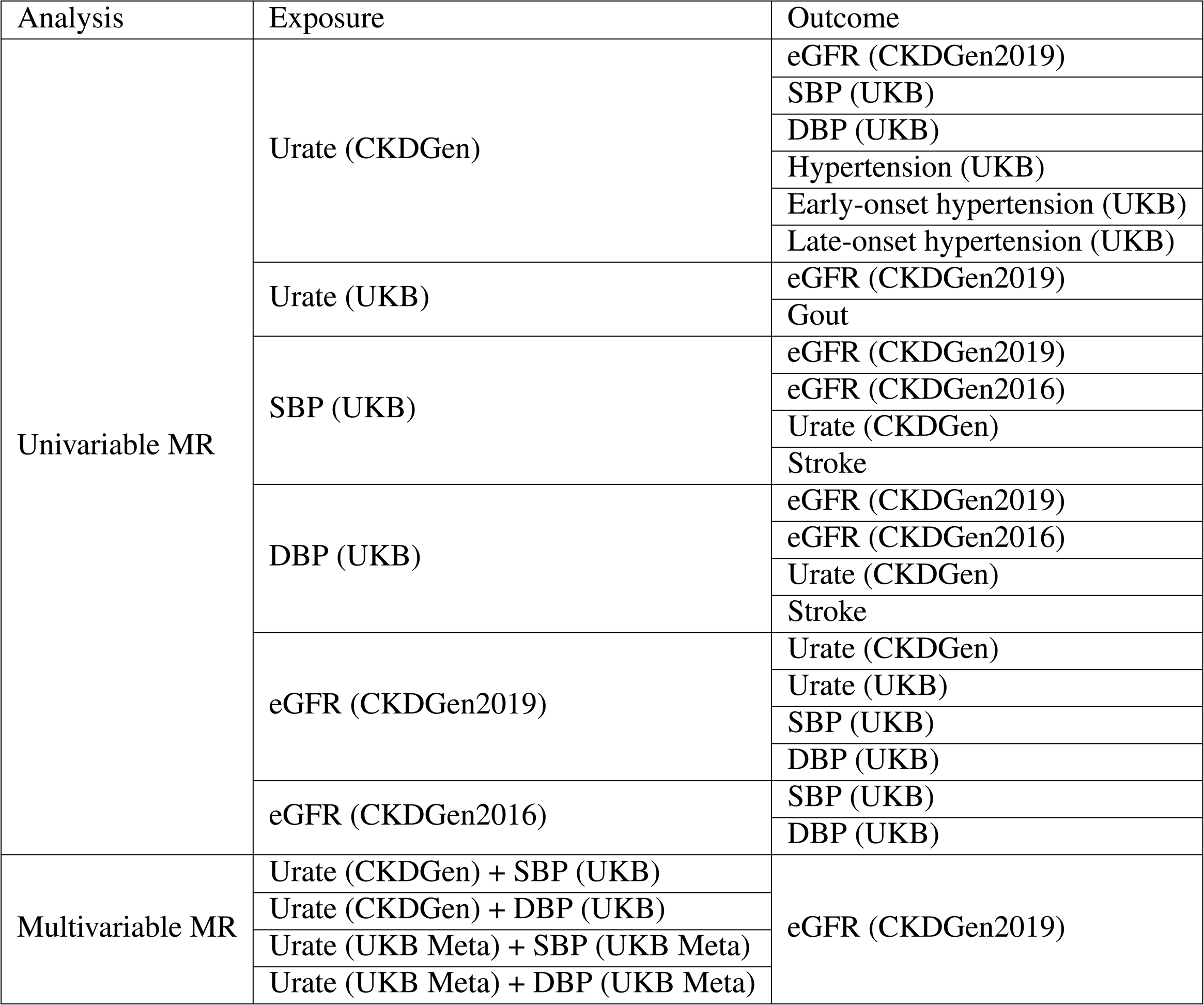
Details of all MR analyses conducted in this study. Each trait was presented as the trait name (its corresponding source). We denote urate data from CKDGen as “Urate (CKDGen)” [18] and from UKB as “Urate (UKB)”, and distinguish two eGFR GWAS from CKDGen as “eGFR (CKDGen2019)” [19] and “eGFR (CKDGen2016)” [20]. To avoid sample overlap between Urate and eGFR, Urate UKB GWAS was used in UVMR while the split-sample method, where the UKB sample was randomly divided into two to conduct GWAS, was used in MVMR. Meta indicated that causal estimates were meta-analyzed from the causal effects of the two subsets. UVMR, univariable Mendelian Randomization; MVMR, multivariable MR; UKB, UK Biobank; SBP and DBP, systolic and diastolic blood pressure; eGFR, estimated glomerular filtration rate; CKDGen, the Chronic Kidney Disease Genetics Consortium.

**Table 2:**
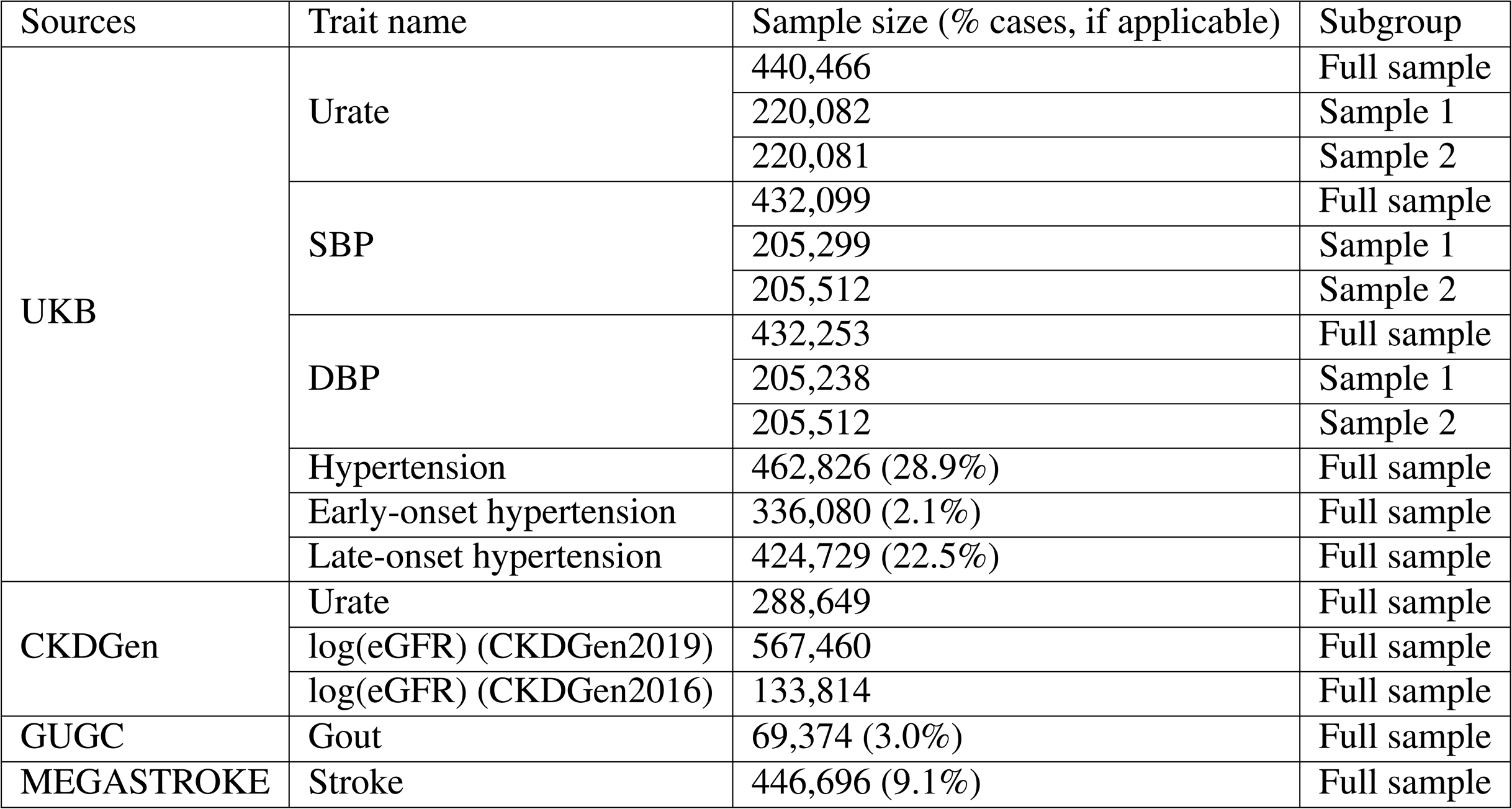
Details of all genome-wide association studies (GWAS) used in our study. All UKB GWAS were newly conducted; “Full sample” indicates that all participants were used; “Sample 1” and “Sample 2” indicate that the split-sample method was used (see details in Methods and materials). UKB, UK Biobank; SBP and DBP, systolic and diastolic blood pressure; CKDGen, the Chronic Kidney Disease Genetics Consortium; eGFR, estimated glomerular filtration rate; GUGC, Global Urate Genetics Consortium.

#### Early- and late-onset hypertension

We examined the causal effects of urate on early-onset, late-onset, and overall hypertension using UVMR. In the literature, the definition of early-onset hypertension varies from *≤* 35 to *≤* 55 years of age [25–30]. Given the UK National Institute for Health and Care Excellence (NICE) makes treatment recommendations based on whether patients are above or below 55 years old [31], we defined the threshold for early-onset and late-onset hypertension as 55 years old. To reduce misclassification, we set a 5-year window before and after the threshold of 55 years old. Therefore, early-onset hypertension was defined as a diagnosis at an age of *≤* 50 years; late-onset hypertension was defined as a diagnosis at an age of > 60 years. To implement the threshold, the year and month of birth of participants, as well as the date that the ICD10 codes I10 and I15 were first reported, were extracted from UKB. We then randomly assigned a day of birth within the birth month to each participant and calculated the approximate age at which participants were diagnosed with hypertension.

#### MVMR

To investigate whether the effects of urate on eGFR are independent of BP, we applied MVMR using summary level urate genetic associations [18] from CKDGen and BP genetic associations from UKB as exposure, with eGFR (CKDGen2019) [19] as the outcome. Genetic instruments in MVMR still need to adhere to the instrumental variable assumptions and must be related to at least one exposure. MVMR allows for the inclusion of multiple exposures and separates the direct causal effects of each exposure in the model. [32]. To determine whether the genetic instruments effectively predict each exposure while considering the presence of the other exposure within the MVMR model, we calculated the conditional F-statistic for each exposure [33]. A conditional F-statistic larger than the arbitrary threshold of 10 indicates that the genetic instruments for MVMR are likely to be strong.

### Data availability

All MR analyses conducted in this study are shown in Table 1 and the details of GWAS data used for MR analyses are in Table 2 (for positive control results, see Supplementary Materials). Our analysis code is available on GitHub (https://github.com/Haotian2020/Urate_Project2024).

## Results

For all results, effect estimates are presented as standard deviation (SD) increase for continuous outcomes and odds ratios (ORs) for binary outcomes with 95% confidence intervals (CIs) and p-value.

### UVMR

#### Genetically predicted urate levels

Genetically predicted higher urate increased SBP and DBP by 0.08 (95% CI: 0.04 to 0.11, p=6.5e-5) and 0.09 (95% CI: 0.05 to 0.14, p=3.5e-5) SD respectively [Figure 1A and Supplementary Table (ST) 3]. The causal effect of urate (CKDGen) on eGFR was −0.10 (95% CI: −0.22 to 0.02, p=0.12), while the effect of urate (UKB) on eGFR was −0.17 (95% CI: −0.24 to −0.09, p=1.6e-5).

**Figure 1:**
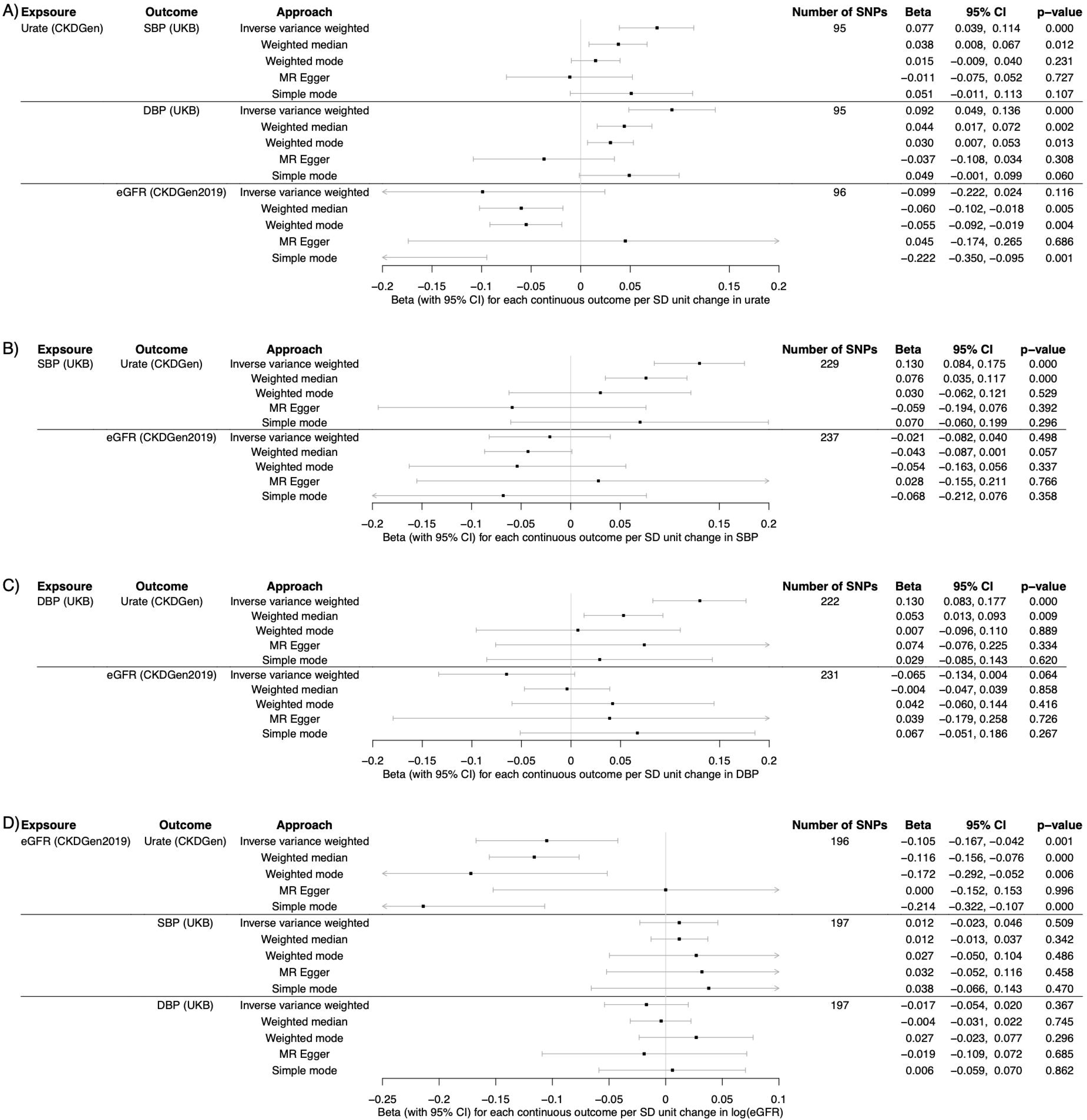
Forest plot of the pairwise bidirectional univariable MR results. Each trait is presented as the trait name (its corresponding source). Estimates of the causal effects of the following exposures: A) urate (CKDGen); B) SBP (UKB); C) DBP (UKB); D) eGFR (CKDGen2019), are presented as SD unit change in outcome per SD unit increase in exposure (eGFR is in the SD unit of log(eGFR)). CI, confidence interval; SBP and DBP, systolic and diastolic blood pressure; eGFR, estimated glomerular filtration rate; MR, Mendelian Randomization.

Genetically increased urate levels led to a higher risk of early-onset, late-onset, and overall hypertension (Figure 2 and ST 7). The odds ratio per SD increase in genetically predicted urate was 1.28 (95% CI: 1.07 to 1.53, p=6.1e-3) for early-onset hypertension, 1.17 (95% CI: 1.10 to 1.26, p=5.4e-6) for late-onset hypertension, and 1.22 (95% CI: 1.12 to 1.32, p=2.5e-6) for overall hypertension.

**Figure 2:**
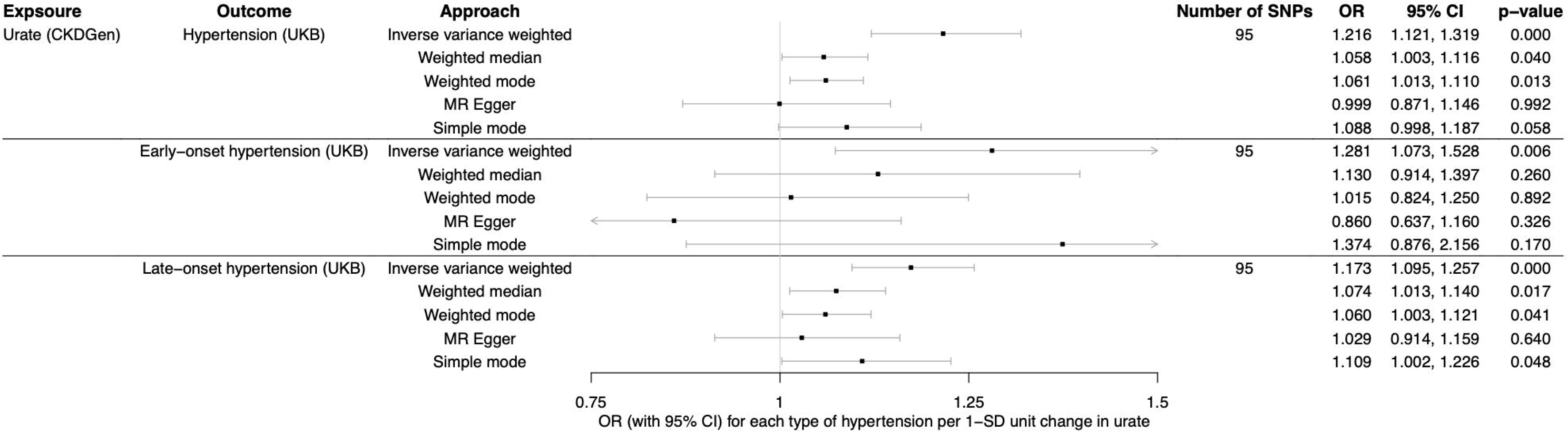
Forest plot of MR results of urate on each type of hypertension. Exposure and outcomes are presented as the trait name (its source name). Estimates of causal effects are presented as OR of each hypertension outcome per SD unit increase in urate (CKDGen). MR, Mendelian randomization; CI, confidence interval. OR, odds ratio.

#### Genetically predicted BP

Genetically predicted higher BP increased urate levels (SBP: beta= 0.13, 95% CI: 0.08 to 0.18, p=2.3e-8; DBP: beta =0.13, 95% CI: 0.08 to 0.18, p=6.5e-8) (Figure 1B and 1 C; ST 3). The causal effects of genetically elevated BP on log(eGFR) were −0.02 (95% CI: −0.08 to 0.04, p=0.50) for SBP and −0.06 (95% CI: −0.13 to 0.004, p=0.64) for DBP.

#### Genetically predicted eGFR levels

Genetically predicted higher log(eGFR) decreased urate levels from both CKDGen (beta=-0.10, 95% CI: −0.17 to −0.04, p=1.0e-3) and UKB (beta=-0.11, 95% CI: −0.18 to −0.04, p=1.1e-3) . The causal effects of genetically predicted log(eGFR) on BP were 0.01 (95% CI: −0.02 to 0.05, p=0.51) for SBP and −0.02 (95% CI: −0.05 to 0.02, p=0.37) for DBP.

#### Sensitivity analyses of UVMR

The genetic instruments used in all UVMR analyses, with their F-statistics and 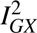 statistics, are shown in ST 2 and 4 respectively. All instruments had F-statistics larger than 10 indicating that they are likely strong instruments. All 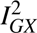 were larger than 98.64% indicating that the NOME assumption is unlikely to have been violated. However, in all UVMR except urate on gout, Cochran’s Q was much higher than its degrees of freedom, indicating the presence of heterogeneity (see ST 3).

The results of pleiotropy tests of UVMR without Steiger filtering are shown in ST 4. There was likely directional pleiotropy in the analysis of urate on early-onset hypertension (intercept=0.016, p=2.2e-3). There was negligible directional pleiotropy in the analysis of urate (CKDGen) on SBP (intercept=0.003, p=1.4e-3) and DBP (intercept=0.005, p=4.1e-5), urate from UKB on eGFR (intercept=-0.004, p=6.7e-3), SBP on urate from CKDGen (intercept=0.003, p=4.0e-3), urate on late-onset hypertension (intercept=0.005, p=1.0e-2) and overall hypertension (intercept=0.008, p=1.0e-3). There was limited evidence of directional pleiotropy for the other main analyses. The weighted median results indicated a similar signal from the pleiotropy tests with evidence of pleiotropy affecting the causal estimate of urate on early-onset hypertension (OR=1.13, 95% CI: 0.91 to 1.40, p=0.26). Other weighted median results showed consistent evidence with the IVW results, consistent with the negligible pleiotropy effect found using MR-Egger, except urate (CK-DGen) on eGFR (weighted median: beta=-0.06, 95% CI: −0.10 to −0.02, p=5.2e-3). The positive control MR analyses confirmed that our instruments could detect known effects (Supplementary Materials and Figure S2).

Although Steiger filtering removed up to 52 SNPs from the UVMR analyses, the causal estimates from IVW remained consistent with the main analyses (ST 7).

#### Sensitivity analyses for eGFR GWAS

The results of bidirectional UVMR between eGFR (CKDGen2016) and BP were consistent with the main analyses involving eGFR (CKDGen2019) and BP, indicating weak evidence of causal effects between eGFR (CKDGen2016) and BP (see Supplementary Materials).

### MVMR

The conditional F-statistics of instruments of each exposure were greater than 10, indicating the genetic instruments in MVMR were strong (ST 8). MVMR provided evidence that increased urate has a causal effect on decreased eGFR, independent of SBP (Figure 3A), and DBP (Figure 3B). Changes in SD log(eGFR) per SD increase in genetically predicted urate were −0.10 (adjusted with SBP, 95% CI: −0.18 to −0.02, p = 1.2e-2) and −0.12 (adjusted with DBP, 95% CI: −0.20 to −0.04, p=4.1e-3). There was little evidence of a causal effect of genetically predicted higher SBP or DBP on eGFR, independent of urate (SBP: beta = −0.03, 95% CI: −0.13 to 0.07, p=0.58; DBP: beta=-0.09, 95% CI: −0.19 to 0.01, p=6.8e-2). All estimates were consistent with the main analyses when repeated using the split-sample method to account for sample overlap (Figure S3).

**Figure 3:**
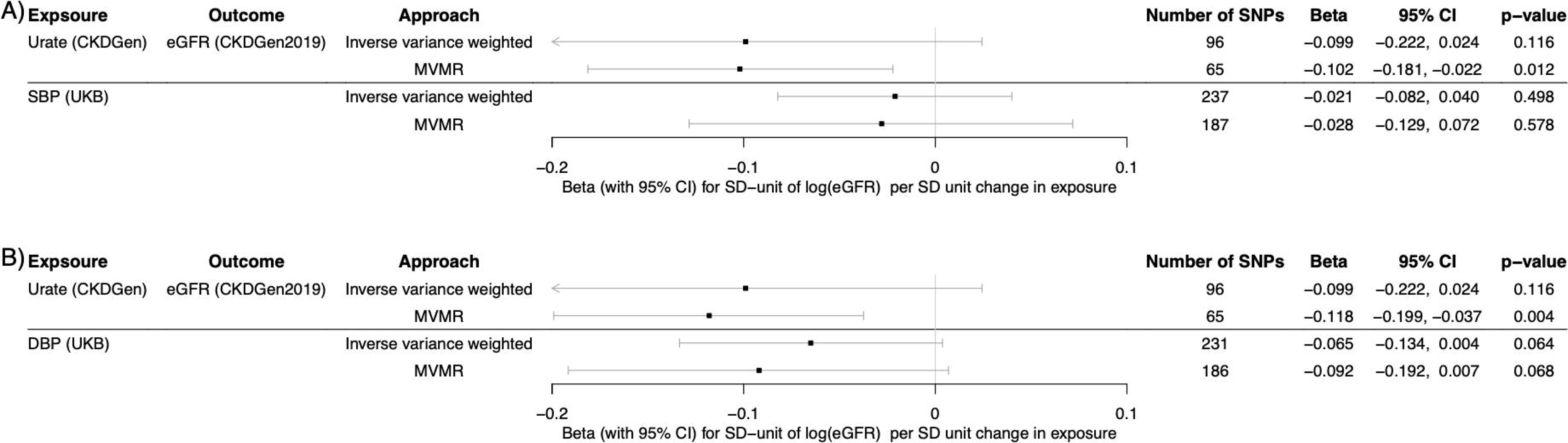
Forest plot of MVMR of urate and BP on eGFR. Each trait is presented as the trait name (its corresponding source). Estimates of causal effects of the two sets of following exposures: A) urate (CKDGen) and SBP (UKB); B) urate (CKDGen) and DBP (UKB), are presented as SD unit change in log(eGFR) per SD unit increase in exposure. CI, confidence interval; SBP and DBP, systolic and diastolic blood pressure; eGFR, glomerular filtration rate estimated from serum creatinine; MVMR, multivariable MR.

## Discussion

We examined the causal relationships between urate, BP, and eGFR by conducting pairwise UVMR and MVMR of urate and BP on eGFR. The UVMR results indicated strong evidence of bidirectional causal effects between urate and BP (SBP and DBP) and between urate and eGFR. There was also evidence of causal effects of urate on early-onset, late-onset, and overall hypertension, but we could not distinguish whether urate had a larger effect on early-onset hypertension. We found inconclusive evidence regarding the causal effects between BP and eGFR. MVMR results indicated that the causal effects of urate on eGFR were independent of BP.

Numerous epidemiological studies have evaluated blood pressure as a risk factor for both the onset and progression of CKD [34–38] and CKD can also arise as a complication of untreated high blood pressure. Previously, a bidirectional MR found strong evidence of the causal effect of higher eGFR on lower SBP and DBP, but not vice versa [39]. This study used eGFR and blood urea nitrogen (BUN) summary statistics to select genetic instruments for eGFR. Given that higher eGFR results in lower BUN, they assumed eGFR instruments relevant to kidney function (i.e., with a positive association with eGFR) should correspondingly have a negative association with BUN. They concluded that higher eGFR reduces BP. However, our study does not support the evidence of a causal effect of eGFR on BP. This could be due to the differences in the instrument selection approach or because the BP GWAS used in the previous study was additionally adjusted for body mass index [40], while we conducted our BP GWAS only adjusting for genotyping chip, sex, and age in UKB. Conducting two-sample MR requires the underlying assumption of a linear relationship between exposure and outcome so we cannot make inferences regarding non-linear relationships between these phenotypes. Due to concerns about the reliability of non-linear MR [41], we did not apply non-linear MR to the analyses between BP and eGFR. The weak evidence of causal effects of eGFR on BP may result from biased estimates due to eGFR overestimation compared to measured GFR using the Chronic Kidney Disease Epidemiology Collaboration (CKD-EPI) equation [42].

Assessing the causal role of urate on kidney function in epidemiological studies is challenging due to the inherent relationship between urate and the kidneys. Urate is primarily excreted by the kidneys, and if kidney function is compromised, there is compensatory but insufficient elimination by the gut [43]. A previous MR study, using 26 SNPs identified from a cohort of 110,347 individuals of European ancestry by the Global Urate Genetics Consortium [44], found limited evidence of a causal relationship between urate and either eGFR or the risk of developing CKD [45]. Another MR study using a genetic urate score indicated hyperuricemia could predict the risk of gout but did not demonstrate predictive power for the development of hypertension or CKD [**?**]. However, these results could reflect the limited number of genetic instruments and outcome sample size used in these studies. Our MR results with larger sample sizes of both urate and other traits showed consistent results with one previous MR study [46], which found serum urate had a causal effect on increased SBP. Additionally, we identified a bidirectional causal relationship, indicating that elevated BP may also lead to increased urate levels. Our study provides further insight by using UVMR with and without Steiger filtering and MVMR, which allows us to estimate the effect of urate, independent of blood pressure, on eGFR and the effect of blood pressure, independent of urate, on eGFR. Moreover, the correlation between urate levels and hypertension diminishes with increasing age and duration of hypertension, suggesting a potential significance of urate in younger individuals with early-onset hypertension [47, 48]. Thus, we conducted a novel MR analysis of urate on early-onset, late-onset, and overall hypertension. While we observed a causal effect of urate on all hypertension types, including some evidence of a directional pleiotropy effect for early-onset hypertension, the wide confidence interval of the effect of urate on early-onset hypertension precludes distinguishing its effect magnitude from that on late-onset hypertension. This is likely due to the sample size as there were only 6,934 cases in the early-onset hypertension

### GWAS

Our study had several strengths. Firstly, we harnessed the power of large sample sizes from UKB and CKDGen to identify strong instruments for our phenotypes of interest and decreased the probability of violation of the MR relevance assumption. Secondly, we proactively addressed sample overlap issues in UVMR and MVMR analyses caused by the fact that both urate and eGFR GWAS were taken from CKDGen. We addressed this by using the GWAS conducted with the full UKB sample and the split-sample method in the UVMR and MVMR sensitivity analyses respectively. Both indicated limited evidence of bias caused by the sample overlap. Thirdly, we conducted all bidirectional MR analyses with Steiger filtering to mitigate potential reverse causality concerns between urate, BP, and eGFR. Lastly, we validated the robustness of our findings through a comprehensive set of MR sensitivity analyses, including approaches robust to pleiotropic effects, such as the weighted median and weighted mode methods in UVMR.

Consideration of limitations is essential when interpreting our results. Although sensitivity analyses have shown that most of the pleiotropy effects in our MR analyses are likely to be balanced, the heterogeneity tests indicate a potential violation of the horizontal pleiotropy assumption. Furthermore, despite a relatively large sample from UKB, the cases of early-onset hypertension were limited because the median age at recruitment in UKB is 58 (minimum: 37; maximum: 73). Thus, the power of our MR analysis of urate on early-onset hypertension was limited, resulting in a wide confidence interval. Finally, we only used European-summary-level data in our analyses, which may limit the generalizability of our findings to other ancestries.

In conclusion, we found bidirectional causal effects between urate and both BP and eGFR in the general population. The effects of urate on eGFR were independent of BP. Our findings suggest that, for the general population, controlling serum urate levels might help to reduce BP and maintain kidney function. Implementing lifestyle modifications or treatment aimed at reducing urate serves as a pragmatic and effective strategy for improving cardiovascular and renal health on a population scale.

## Supporting information

Not applicable

## Data Availability

The summary-level data from CKDGen are publicly available (http://ckdgen.imbi.uni-freiburg.de/) and OpenGWAS (https://gwas.mrcieu.ac.uk/). The UK Biobank individual-level data can be accessed through an application (https://www.ukbiobank.ac.uk/).

https://gwas.mrcieu.ac.uk/

https://www.ukbiobank.ac.uk/

## Disclosure

TRG receives funding from Biogen and GSK for unrelated research.

## Data Statement

The summary-level data from CKDGen are publicly available (http://ckdgen.imbi.uni-freiburg. de/ and OpenGWAS https://gwas.mrcieu.ac.uk/ [49]). The UK Biobank individual-level data can be accessed through an application (https://www.ukbiobank.ac.uk/).

## Acknowledgments

The authors thank the CKDGen consortium (https://ckdgen.imbi.uni-freiburg.de) for publicly sharing the summary-level data. This research has been conducted using the UK Biobank Resource under Application No. 15825. This work was supported by the UK Medical Research Council Integrative Epidemiology Unit (MC_UU_00011/4 and MC_UU_00032/03).

## Author Contributions

HT conducted the analyses and drafted the manuscript. VMW and TRG equally contributed to the supervision and manuscript review. All authors approved the final version for submission.

## Notes

### Competing Interest Statement

The authors have declared no competing interest.

### Author Declarations

UK Biobank has approval from the North West Multi-centre Research Ethics Committee (MREC) as a Research Tissue Bank (RTB) approval. This research has been conducted with the UK Biobank Resource under application number 15825.

### Summary of Updates

1. Results section titles updated 2. STROBE-MR file updated 3. Figures updated in TIFF format 4. Discussion words cut

