## Supplementary Materials for "Evaluating the causal relationships between urate, blood pressure, and kidney function in the general population: a two-sample Mendelian Randomization study"

[*]Medical Research Council Integrative Epidemiology Unit, Bristol Medical School, University of Bristol, Bristol, United Kingdom

### Supplementary Methods

#### GWAS in UKB

UKB, a large population-based cohort, recruited over 500,000 individuals and collected extensive phenotypic and genotypic data [1]. We used the Medical Research Council Integrative Epidemiology Unit (MRC IEU) UKB GWAS pipeline to perform our GWAS [2]. We applied the linear mixed modeling approach, BOLT-LMM, to account for population stratification and relatedness. We included genotyping chip, sex, and age as covariates in our models.

We conducted urate, SBP, and DBP GWAS using the full UKB sample. For each continuous trait, data were cleaned by removing extreme values, defined as values more than four standard deviations (SDs) from the mean. The urate GWAS with the UKB full sample was used for a sensitivity analysis to avoid sample overlap, which can cause bias in inverse-variance weighted (IVW) UVMR estimates [3], as both urate [4] and eGFR [5] were from CKDGen. We also randomly divided the participants into two halves. For both subsets, we repeated the urate, SBP, and DBP GWAS. This split-sample approach provided GWAS results in independent samples to address sample overlap concerns in the MVMR analyses.

We also conducted early-onset, late-onset, and overall hypertension GWAS. Hypertension was defined as participants with ICD10 codes I10 or I15 (primary and/or secondary hypertension respectively).In the literature, the definition of early-onset hypertension varies from $\leq 35$ to $\leq 55$ years of age [6–11]. Given the UK National Institute for Health and Care Excellence (NICE) makes treatment recommendations based on whether patients are above or below 55 years old [12], we defined the threshold for early-onset and late-onset hypertension as 55 years old. To reduce misclassification, we excluded individuals diagnosed with hypertension in the 5-years before or after the threshold age. Therefore, early-onset hypertension was defined as a diagnosis at an age of $\leq 50$ years; late-onset hypertension was defined as a diagnosis at an age of $> 60$ years. To implement the threshold, the year and month of birth of participants and the first date of a relevant ICD10 code were extracted from UKB. We then randomly assigned a day of birth within the birth month to each participant and calculated the approximate age at which participants were diagnosed with hypertension.

#### MR assumptions

As shown in Figure S1, MR relies on the validity of three core assumptions [13]:

1: The instrument is strongly associated with exposure (Relevance).

2: The instrument and the outcome have no common causes (Independence).

3: The instrument is associated with the outcome via the exposure only (Exclusion restriction).

#### Estimation of trait standard deviation (SD)

The estimate of the SD of log(eGFR) in Wuttke *et al.* (2019) was based on the approach taken by a previous publication [14], where they used the SD of log(eGFR) from European-ancestry participants of the Atherosclerosis Risk in Communities Study (ARIC) study to present the overall SD of the meta-analyses. The SD of the log(eGFR residuals) was estimated to be 0.13. The SD of log(eGFR) in Pattaro *et al.* 2016 was 0.24, obtained from the IEU open GWAS database (ID: ieu-a-1105) [15].

We obtained the SD of urate from CKDGen by calculating the weighted average, where we assigned each meta-analyzed cohort-specific sample size as the weighting factor [4]. The SD of urate from CKDGen was 1.5 mg/dL. The SD of urate, SBP, and DBP in UK Biobank was 1.3 mg/dL, 19.5 mmHg, and 10.6 mmHg respectively.

#### Sensitivity analyses

##### Sensitivity analyses for UVMR

By assuming that all exposure SNPs are valid instrumental variables, the IVW method combines the effects of all these SNPs to obtain an overall weighted effect [16]. However, the validity of MR estimates depends on three assumptions (Figure S1). To evaluate the robustness of our findings [13], we performed different sensitivity analyses, including MR-Egger [17], weighted median [18], weighted mode, simple mode [19] and Steiger filtering [20]. When balanced pleiotropic effects are present, the MR-Egger method exhibits resilience by permitting the intercept in the regression of the SNP-outcome association against the SNP-exposure association to be non-zero. The intercept term in the MR-Egger result can be used as an assessment of directional pleiotropy, with the intercept term being interpreted as a measure of the directional pleiotropy present (pleiotropy tests) [17]. MR-Egger makes the 'NO Measurement Error' (NOME) assumption [21, 22]. We calculated the $I^2_{GX}$ statistics to assess any violation of the NOME assumption. When taken as an estimate of the attenuation bias, an $I^2_{GX}$ statistic higher than 90% corresponds to less than 10% relative bias towards the null [21]. The weighted median method can provide reliable results even when a maximum of 50% of the instruments are not valid [18]. The mode method assumes that instruments from the largest subset are valid and identify the same true causal effect [19]. Finally, we applied Steiger filtering to ensure that each genetic instrument had a stronger association with the exposure than with the outcome, which minimized the risk of reverse causation in bidirectional MR [20].

We applied Cochran's Q-test to assess heterogeneity in each MR analysis by examining the variability in the causal effects of each genetic instrument. When Q significantly exceeds its degrees of freedom (which is calculated as the number of SNPs minus 1), it indicates the presence of heterogeneity [23, 24].

To address the sample overlap issue that both the urate and eGFR GWAS from CKDGen used many of the same individuals, we performed a urate GWAS using UKB data and repeated our UVMR analyses to assess the causal relationship between urate and eGFR in non-overlapping samples.

Additionally, to validate the instruments for urate, SBP, and DBP, we performed three positive control MR analyses. These were urate on gout, SBP on stroke, and DBP on stroke (see Supplementary Materials).

##### Sensitivity analyses for MVMR

Due to the sample overlap problem in which urate [4] and eGFR [5] GWAS were both from CKDGen, we used urate from UKB as a sensitivity analysis. We applied the split-sample method for urate, SBP, and DBP to ensure non-overlapping samples in each MVMR analysis. We then conducted MVMR analyses using each split sample, followed by a meta-analysis using a fixed-effect model to obtain a single estimate. The conditional F-statistic of each exposure was calculated as the mean conditional F-statistic of the meta-analyzed MR analyses.

**Sensitivity analyses for eGFR GWAS**

Multiple eGFR GWAS datasets are available from CKDGen [5, 25, 26]. To further assess the causal relationship between BP and eGFR as well as address the sample overlap problem in MR analyses, we used eGFR GWAS [25] (OpenGWAS [15] ID: ieu-a-1105) for sensitivity analyses between eGFR and BP. Additionally, we used BP instruments to compare their SNP effect on log(eGFR) from two eGFR GWAS [5, 25] (see Supplementary Materials).

#### Positive control: Univariable MR (UVMR) of urate on gout and BP on stroke

To validate our GWAS of urate, SBP, and DBP, we examined the effect of urate on gout, as well as the effects of SBP and DBP on stroke by conducting two-sample MR analyses. The urate instruments were extracted from the urate, SBP, and DBP GWAS conducted in UKB (see Method and Materials). The gout GWAS summary statistics were obtained from a meta-analysis study of 14 studies, including 2,115 cases and 67,259 controls [27] (OpenGWAS [15] ID: ebi-a-GCST001790). We obtained the stroke GWAS from the MEGASTROKE consortium in the European population, including 40,585 cases and 406,111 controls [28].

#### Bidirectional UVMR between BP and eGFR (CKDGen2016)

To validate the bidirectional MR results between BP and eGFR, we did additional bidirectional UVMR analyses between BP and eGFR, by using a previous eGFR GWAS [25]. We label two eGFR GWAS as "eGFR (CKDGen2019)" and "eGFR (CKDGen2016)" representing Wuttke *et al.* (2019) and Pattaro *et al.* (2016) respectively. The BP instruments were extracted from the SBP and DBP GWAS conducted in UKB (see Method and Materials). The previous eGFR GWAS summary statistics were obtained from OpenGWAS [15] (ID: ieu-a-1105), including 133,814 European population. The instruments of the previous eGFR GWAS were the same as described in Instrument selection. To compare the difference in SNP effects between two eGFR GWAS, we generated the scatter plots by using the BP instruments, with the x and y axes representing the SNP effects on eGFR (CKDGen2019) [5] and eGFR (CKDGen2016) [25] respectively.

#### Software

Linkage disequilibrium (LD) clumping for genetic instruments was conducted using the `ieugwasr` [29] R package (`https://mrcieu.github.io/ieugwasr/`). All UVMR and MVMR analyses were performed with the `TwoSampleMR` [30] R package (`https://github.com/MRCIEU/TwoSampleMR`). Conditional F-statistics for exposures in MVMR were calculated using the `MVMR` [31] R package (`https://github.com/WSpiller/MVMR`). Forest plots were generated using the `forestplot` R package (`https://github.com/gforge/forestplot`).

### Supplementary Results

#### Baseline characteristics

As shown in Supplementary Table (ST) 1A, among the 460,727 participants in the UK Biobank (UKB), the median age was 58.0 years, with 45.7% being male. The median urate level was 5.1 mg/dL, and the median SBP and DBP were 138.0 and 82.0 mmHg. We identified 21.8% of participants who had been diagnosed with hypertension and 1.5% with CKD, as shown in ST 1A. We estimated the median eGFR in UKB participants by using serum creatinine with the Chronic Kidney Disease Epidemiology Collaboration (CKD-EPI) Equation [32]. UKB participants with the ICD-10 code N18 were considered as CKD cases and the rest of them were considered as control.

In the eGFR 2019 GWAS [5] from the CKDGen Consortium, which included 567,460 participants, the median age was 50.1, and 48.2% were male. The median eGFR in this sample was 91.4 ml/min per 1.73 $m^2$ (ST 1A). For the meta-analysis of the GWAS of serum urate levels within the CKDGen Consortium, which involved 288,649 participants, the mean age was 57.9 years, with 61.9% being male. The mean urate level was 5.5 mg/dL (ST 1B).

#### Instrument Selection

From CKDGen, we extracted 201 SNPs for eGFR (CKDGen2019) [5], 47 SNPs for eGFR (CKDGen2016) [25] and 96 SNPs for urate (CKDGen) [4]. From the UKB sample, we obtained 322

SNPs from the GWAS of the full urate sample, 164 SNPs from the GWAS of urate in sample 1, and 171 SNPs from the GWAS of urate in sample 2. Similarly, for SBP, we obtained 264, 91, and 80 SNPs from the full sample, sample 1, and sample 2 respectively; for DBP, we obtained 254, 86, and 77 SNPs from the full sample, sample 1, and sample 2 respectively.

#### Main analyses

##### Positive control UVMR results

UVMR positive control analyses with and without Steiger filtering are provided in ST 7C and 7D. There was consistent strong evidence among all MR methods that genetically predicted urate level causally increased the risk of gout with an OR per 1-SD unit increase in urate of 4.77 [Inverse variance weighted (IVW) method: 95% CI: 3.82 to 5.97, p = 5.63e-43]. As shown in Figure S2, there was strong evidence that genetically predicted BP level causally increased the risk of stroke (for SBP: OR=1.76, 95% CI: 1.58 to 1.96, p=7.4e-25; for DBP: OR=1.55, 95% CI: 1.39 to 1.72, p=1.5e-15).

##### Bidirectional UVMR between BP and eGFR (CKDGen2016)

The bidirectional UVMR analyses between BP and eGFR (CKDGen2016) with and without Steiger filtering are provided in ST 5 and 6. There was little evidence of the causal effects of genetically predicted higher BP levels on decreased log(eGFR) (SBP: beta=-0.04, 95% CI: -0.13 to 0.05, p=0.40; DBP: beta=-0.07, 95% CI: -0.18 to 0.03, p=0.18), as well as of genetically predicted higher log(eGFR) on SBP (beta=0.01, 95% CI: -0.04 to 0.06, p=0.71) or DBP (beta=-0.03, 95% CI: -0.09 to 0.02, p=0.21). These results are consistent with our main MR analyses between BP and eGFR (CKDGen2019).

#### Sensitivity analyses

Cochran's Q of SBP and DBP on stroke as well as between BP and eGFR (CKDGen2016) were much higher than their degree of freedom, showing the presence of heterogeneity while Q of urate on gout is slightly higher than its degree of freedom (p=0.43) (ST 3). The F-statistics of urate, SBP, DBP, and eGFR (CKDGen2016) instruments were larger than 10, indicating that the genetic instruments were strong. The $I^2_{GX}$ were larger than 98.17% suggesting that the MR-Egger estimates were unlikely to be biased.

The pleiotropy test showed that there was strong evidence for some directional pleiotropy effects in the analysis of urate on gout (intercept=-0.016, p=6.88e-5). Limited evidence of directional pleiotropy effects was observed for SBP and DBP on stroke (SBP: intercept=-0.003, p=0.28; DBP: intercept=-0.004, p=0.26) and between BP and eGFR (CKDGen2016) (SBP on eGFR: intercept=-0.001, p=0.60; DBP on eGFR: intercept=-0.003, p=0.28; eGFR on SBP: intercept=0.003, p=0.41; eGFR on DBP: intercept=0.002, p=0.59). All sensitivity analyses for the positive control MR were consistent with the main estimates for the positive control MR, indicating instruments obtained from the novel GWAS conducted in UKB were appropriate for our analysis. All sensitivity analyses for bidirectional MR between BP and eGFR (CKDGen2016) have wide confidence intervals crossing the null and indicating insufficient statistical power. All Steiger filtering results showed consistent results without Steiger filtering.

In Figure S4, we observed attenuated effects on log(eGFR) for both SBP (top panel) and DBP (bottom panel) instruments in eGFR (CKDGen2019) compared to eGFR (CKDGen2016), notably in log(eGFR) unit (left panel), and less prominently in SD unit of log(eGFR) (right panel). This consistency aligns with MR findings where point estimates of BP on eGFR (CKDGen2019) were smaller than those on eGFR (CKDGen2016), while the confidence intervals of BP on eGFR (CKDGen2019) were narrower than those on eGFR (CKDGen2016), as eGFR (CKDGen2019) have larger sample size.

13. Lawlor DA, Harbord RM, Sterne JAC, Timpson N, Davey-Smith G. Mendelian randomization: Using genes as instruments for making causal inferences in epidemiology. *Statistics in*

*Medicine.* 2008;27(8):1133–1163.

21. Bowden J, Del Greco M. F, Minelli C, Davey-Smith G, Sheehan NA, Thompson JR. Assessing the suitability of summary data for two-sample Mendelian randomization analyses using MR-Egger regression: the role of the I2 statistic. *International Journal of Epidemiology.* 2016:dyw220.

### Supplementary Figures

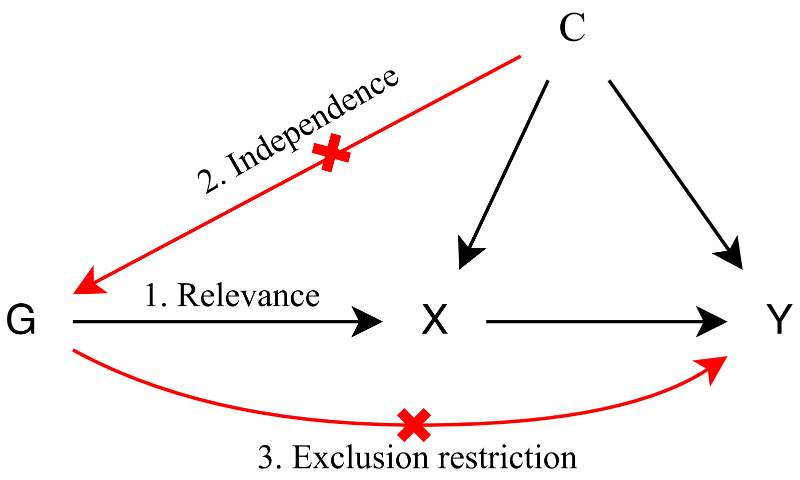


Figure S1: The three core assumptions for MR (relevance, independence, and exclusion restriction). MR, Mendelian Randomization; G, genetic instruments; X, exposure; Y, outcome; C, confounder.

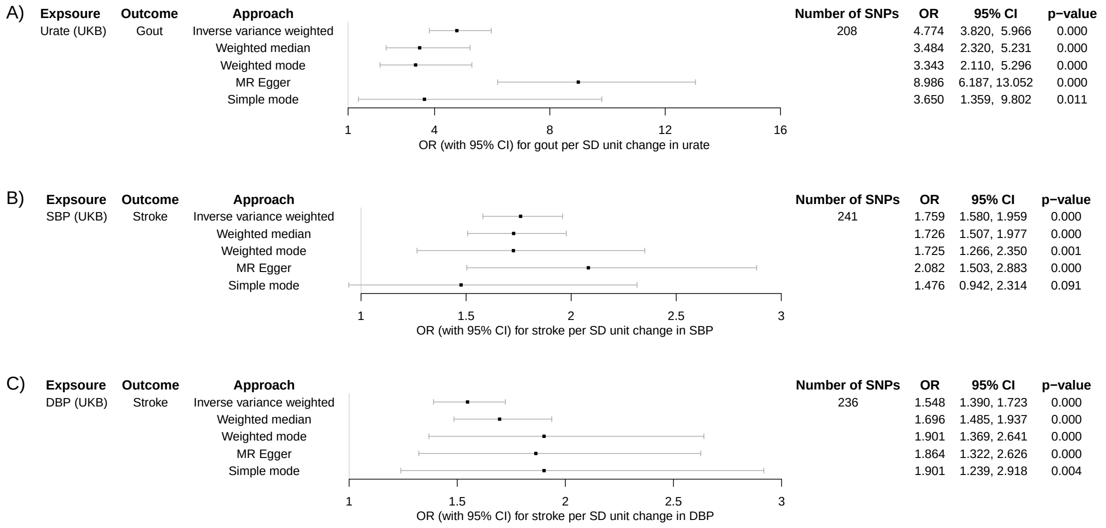


Figure S2: The forest plot of positive control MR analyses for A) urate on gout; B) SBP on stroke; and C) DBP on stroke. MR, Mendelian Randomization; UKB, UK Biobank; SBP and DBP, systolic and diastolic blood pressure; CI, confidence interval.

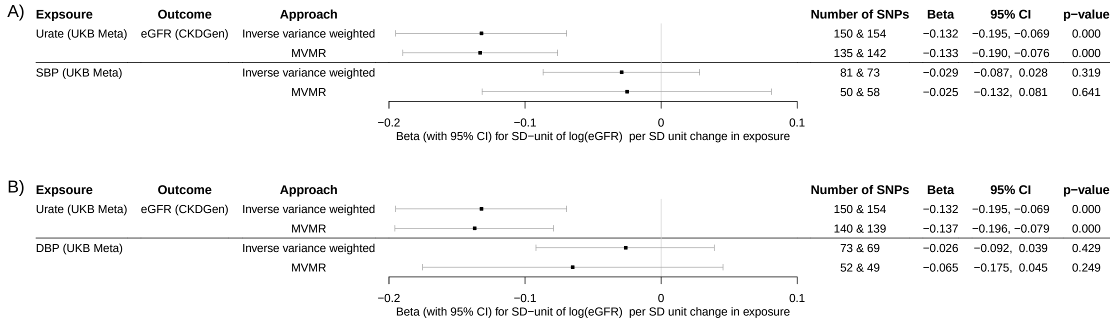


Figure S3: The forest plot of MVMR sensitivity analysis. To avoid the sample overlap problem, as both the urate and eGFR GWAS were from CKDGen, we applied the sample-split method. We therefore conducted urate, SBP, and DBP GWAS in UKB by randomly dividing the full sample into two samples. The number of SNPs column indicates the number of SNPs included in each MR before meta-analysis. Meta indicates results from the meta-analysis. MR, Mendelian Randomization; MVMR, multivariable MR; UKB, UK Biobank; SBP and DBP, systolic and diastolic blood pressure; CI, confidence interval; eGFR, estimated glomerular filtration rate; CKDGen, the Chronic Kidney Disease Genetics Consortium.

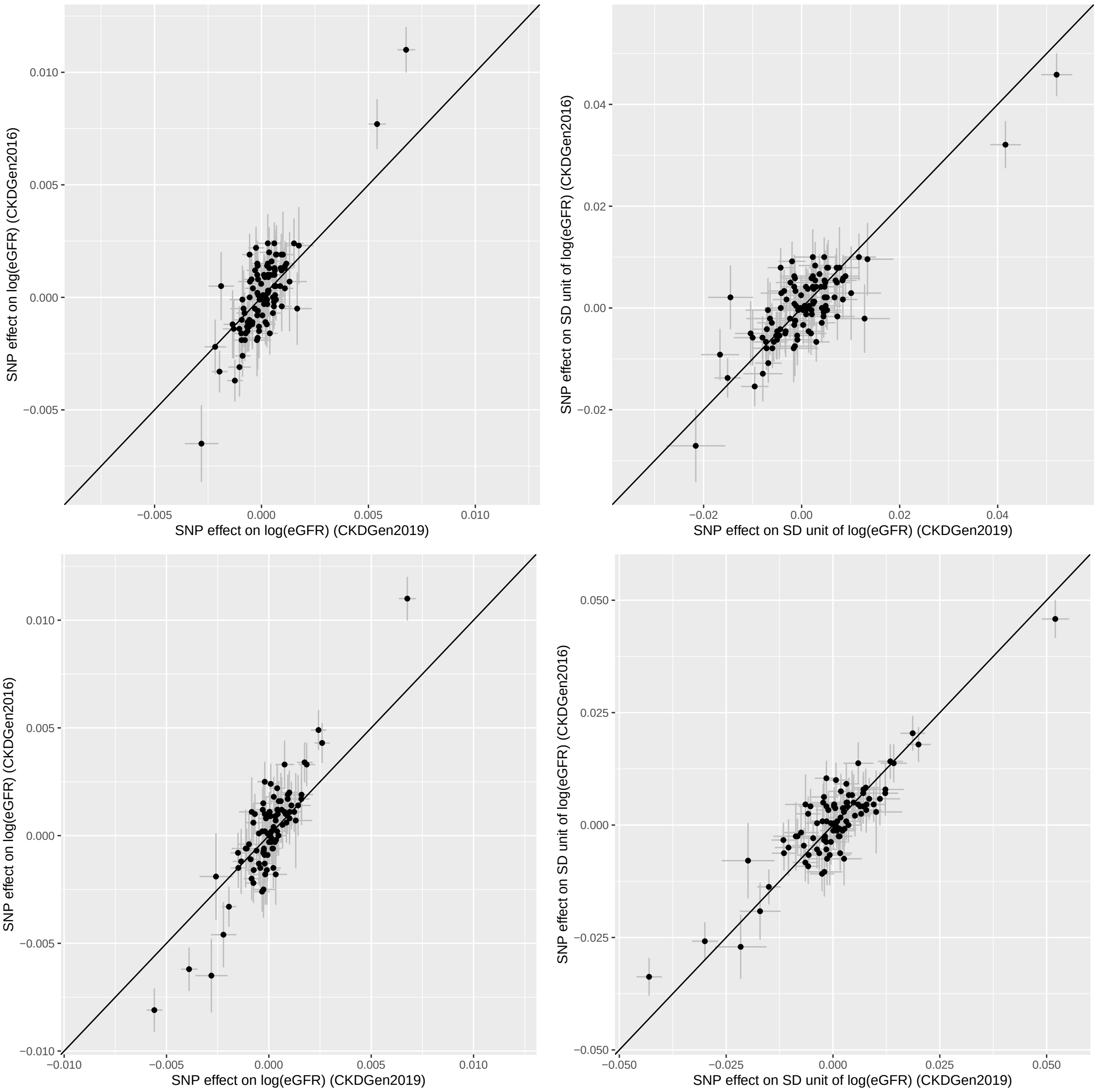


Figure S4: The scatter plots of eGFR GWAS comparison. We compared two eGFR GWAS from CKDGen [5, 25], respectively labeled as "eGFR (CKDGen2016)" and "eGFR (CKDGen2019)", by using SBP (top panel) and DBP (bottom panel) instruments from UKB. Within each panel, we compared the SNP effect on eGFR using log(eGFR) unit (left panel) and SD unit of log(eGFR) (right panel). SBP and DBP, systolic and diastolic blood pressure; UKB, UK Biobank; eGFR, estimated glomerular filtration rate; CKDGen, the Chronic Kidney Disease Genetics Consortium.
